# Automated Deep Brain Stimulation programming based on electrode location – a randomized, cross-over trial using a data-driven algorithm

**DOI:** 10.1101/2022.04.08.22272471

**Authors:** Jan Roediger, Johannes Achtzehn, Johannes L. Busch, Till A. Dembek, Anna-Pauline Krämer, Gerd-Helge Schneider, Patricia Krause, Andreas Horn, Andrea A. Kühn

**Author notes:** **Corresponding Author** Jan Roediger, Movement Disorders and Neuromodulation Unit, Department of Neurology, Charité University Medicine, Charitéplatz 1, Berlin, 10117, Germany.

## Abstract

**Background:** Deep brain stimulation (DBS) of the subthalamic nucleus (STN) is highly effective in controlling motor symptoms in patients with Parkinson’s Disease (PD). However, correct selection of stimulation parameters is pivotal to treatment success and currently follows a time-consuming and demanding trial-and-error process. We conducted a double-blind, randomized, cross-over, non-inferiority trial to assess treatment effects of stimulation parameters suggested by a recently published algorithm (*StimFit*) based on neuroimaging data.

**Methods:** The trial was carried out at Charité – Universitätsmedizin, Berlin, Germany and enrolled 35 PD patients treated with directional octopolar electrodes targeted at the STN. All patients had undergone DBS programming according to our centers standard of care (SoC) treatment before study recruitment. Based on perioperative imaging data DBS electrodes were reconstructed and *StimFit* was applied to suggest optimal stimulation settings. Patients underwent motor assessments using MDS-UPDRS-III during OFF-medication and in OFF-and ON-stimulation states under both conditions, *StimFit* and SoC parameter settings that were double blinded and randomized in a 1:1 ratio. The primary endpoint of this study was the absolute mean difference between MDS-UPDRS-III scores under *StimFit* and SoC stimulation, with a non-inferiority margin of five points.

**Findings:** STN DBS resulted in mean MDS-UPDRS-III improvements of 48 % for SoC and 43 % with *StimFit* as compared to OFF-stimulation condition. The mean difference between MDS-UPDRS-III scores under *StimFit* and SoC stimulation was not significant (1.6 points), and non-inferiority was established. In six patients (17 %) initial programming of *StimFit* settings resulted in acute side-effects and amplitudes were reduced until side-effects disappeared.

**Interpretation:** Automated data-driven algorithms can predict stimulation parameters which lead to motor symptom control comparable to standard of care treatment. This approach could significantly decrease the time necessary to obtain optimal treatment parameters thereby fostering the design of more complex DBS electrodes. Long-term data including effects on quality of life require further investigation.

## INTRODUCTION

Deep Brain Stimulation (DBS) of the subthalamic nucleus (STN) is a well-established treatment option for advanced Parkinson’s Disease (PD), improving motor symptoms, quality of life and allowing to reduce dopaminergic medication.^1-3^ Yet, treatment success depends on the correct selection of stimulation parameters, which includes adaptation of amplitude, stimulation frequency, pulse-width, and the relative distribution of electric current across contacts.^4^ Currently, strategies to optimize these parameters are exclusively based on clinical testing and require highly trained medical personnel to iteratively adjust DBS settings in response to therapeutic or adverse effects.^5^ Typically, this process is initiated by a monopolar review for contact selection and amplitude adjustment, the two most important parameters for effective DBS.^6^ Other parameters are often refined at later stages according to patients’ symptomatic profile and treatment response. This procedure, however, is highly time consuming and impeded by multiple factors including a delayed response to parameter adjustments, symptom fluctuations and patient fatigue. Hence, only a fraction of the vast number of parameter combinations can be evaluated in this manner imposing the risk of selecting suboptimal settings even despite multiple thorough and time-consuming programming sessions. This problem has been aggravated with the introduction of directional electrodes, which allow for a more flexible shaping of the electric field but come at the cost of further inflating the number of possible stimulation settings.^7^

To utilize the full therapeutic potential of modern DBS systems, data-driven algorithms could guide DBS programming by suggesting a subset of stimulation parameters.^8,9^ Electrode localization represents a promising input feature for such an approach since numerous studies have established a link to therapeutic or adverse DBS-effects across various stimulation targets and diseases.^10-15^ This seems especially feasible, since electrodes can be reconstructed from routinely acquired perioperative neuroimaging data allowing for a potential implementation in clinical routine without the need of additional data acquisition or equipment.^16-19^ Commercial software which can provide visual feedback for neurologists in relation to patients’ individual anatomy to aid clinical programming procedures is already available and first prospective applications indicate a potential benefit by reducing the time needed for clinical programming.^20-23^ Despite these advantages image-guided optimization of DBS parameters remains challenging for two reasons. First, to derive optimal stimulation settings iterative adjustments of DBS parameters need to be conducted manually within the software. Although this allows for a much faster probing of different settings, this approach still does not solve the initial problem considering >10^10^ combinatorial possibilities to distribute electric current in octopolar electrodes. Second, optimization objectives are unknown. Decision making is currently based on visualizations of simplified volumetric estimates of neuronal activation (VTA) and their overlap with anatomical regions. However, stimulation targets as well as regions of avoidance are not clearly defined and many therapeutic as well as adverse effects are likely being mediated by a complex interplay of concurrent modulation of multiple white matter bundles.^24^ Hence, decisions are driven (and potentially mislead) by the programmers’ individual understanding of the interplay between anatomy and function.

Aiming at overcoming those limitations we recently developed and retrospectively validated *StimFit*, a data-driven algorithm capable of suggesting optimal stimulation parameters in PD patients treated with STN-DBS based on electrode location in a fully automated fashion.^25^ The model was trained on a large set of standardized monopolar review data to predict therapeutic effects, as well as side-effect probabilities of different stimulation settings. An optimization algorithm was then implemented to find the stimulation setting which would maximize the therapeutic benefit while being constrained by a user-defined side-effect threshold. In the present randomized double-blind cross-over study we now prospectively assess the clinical effects of suggestions made by *StimFit* to the ones derived during standard of care (SoC) programming strategies.

## Methods

### Study design

This double-blind 2 × 2 cross-over non-inferiority trial was designed to evaluate the clinical effects of DBS settings suggested by *StimFit*, a fully automated, data-driven algorithm based on neuroimaging data ^25^ and to compare results to standard of care (SoC). The study was carried out at Charité – Universitätsmedizin, Berlin, Germany and approved by the local ethics committee (EA2/117/19). The study was registered at the German Register for Clinical Trials (https://www.drks.de, Study-ID: DRKS00023115).

### Participants

35 PD patients who underwent STN-DBS surgery with directional octopolar electrodes at Charité were included in the study. Inclusion criteria were the diagnosis of PD according to the British Parkinson’s Disease Society Brain Bank without severe cognitive impairment, neuropsychiatric symptoms or severe cerebral atrophy and the ability to undergo overnight withdrawal of dopaminergic medication. DBS surgeries had to be carried out between three months and three years before recruitment without any major surgical complications like bleedings, infections of the DBS system or re-implantation of DBS electrodes. All patients gave informed written consent.

### Standard of care (SoC)

SoC programming was conducted before enrollment according to the standard postoperative pipeline at our center. This included initial titration of stimulation amplitudes postoperatively and individualized programming approximately three months after surgery. Here, monopolar review examinations were conducted to identify therapeutic windows under cathodal stimulation and DBS settings were adjusted along with the dopaminergic medication over the course of several days during in-patient stays.^26^ Additionally, patients underwent multiple follow-up adjustments at specialized in-and outpatient facilities before study participation (**Suppl. Fig. 1**). All DBS adjustments were carried out by experienced movement disorders specialists who were not involved in any other aspects of the study.

### StimFit setting

Electrodes were reconstructed and normalized to Montreal Neurological Institute (MNI) coordinate system based on multimodal preoperative Magnetic Resonance Imaging (MRI) and postoperative Computed Tomography (CT) using the default Lead-DBS pipeline as described in the original publication of the algorithm.^17,25^ Algorithmic suggestions of DBS parameters were then computed using *StimFit*. Briefly, the model predicts motor improvements and side-effect probabilities of stimulation settings based on electrode reconstructions and simulations of the electric field in the target region. To identify which contact selection and stimulation amplitude would lead to the maximal therapeutic benefit an optimizer iterated through different stimulation settings until the model converged to a final solution (**Figure 1**). To constrain stimulation parameters *StimFit* required to specify a maximum side-effect probability, which was set to 20% (50% within the first four patients). Since the occurrence of tremor varied across patients, tremor was excluded from the prediction, i.e., the model optimized DBS parameters exclusively based on akinetic-rigid symptoms. Computational time to obtain DBS parameter suggestions for both hemispheres was approximately 50 minutes on a local workstation. The resulting stimulation parameters will be referred to as *StimFit* settings.

**Figure 1:**
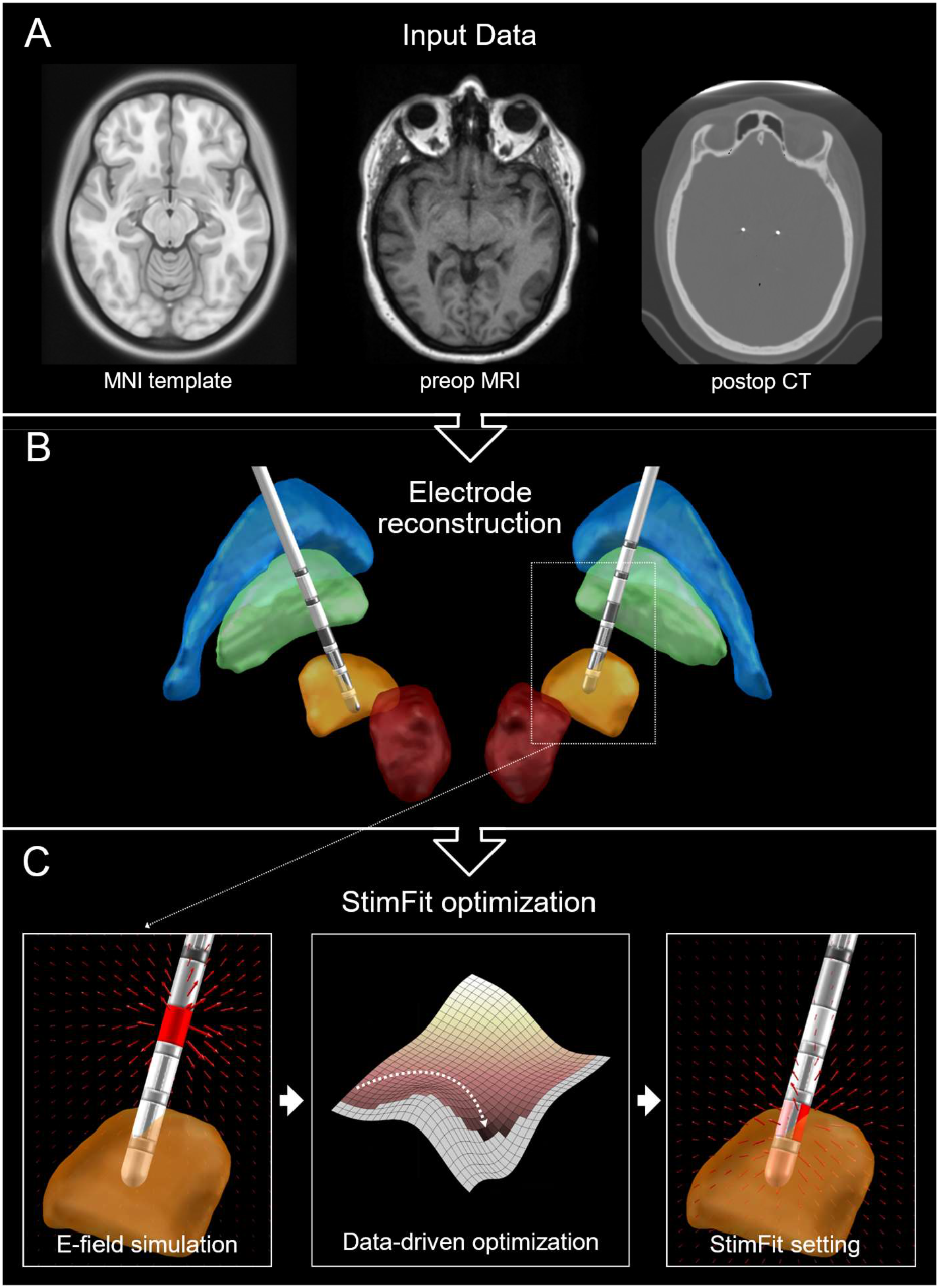
Image-based optimization of DBS parameters. **A & B:** Perioperative neuroimaging data (preoperative MRI and postoperative CT) is processed using the Lead-DBS pipeline to reconstruct DBS electrodes in MNI-space. **C:** Based on electric field simulations *StimFit* predicts clinical effects of different stimulation settings. A nonlinear optimization algorithm iteratively explores the predicted outcomes to converge to a final optimal solution (*StimFit* setting); MNI = Montreal Neurological Institute; MRI = magnetic resonance imaging; CT = computed tomography

### Study protocol

Examinations were performed within one day after overnight withdrawal of dopaminergic medication according to the study protocol (Figure 2). In the preparation phase contact impedances were measured to confirm integrity of the DBS system and to estimate energy efficiency of stimulation settings (see **suppl. material**). *StimFit* settings were activated and assigned to a stimulation program on the pulse generator. In case any acute permanent side-effects occurred, stimulation amplitudes were reduced until side-effects disappeared. Stimulation was then switched off and motor impairment was evaluated after a 45-minute wash-out period using part III of the Movement Disorder Society-Sponsored Revision of the Unified Parkinson’s Disease Rating Scale (MDS-UPDRS-III). Both SoC and *StimFit* ON-stimulation conditions were then evaluated after 45-minutes wash-in periods in randomized order. All ratings were carried out by the same rater (JR). Finally, patients were asked to guess the randomization sequence and to self-assess the effects of both stimulation conditions.

**Figure 2:**
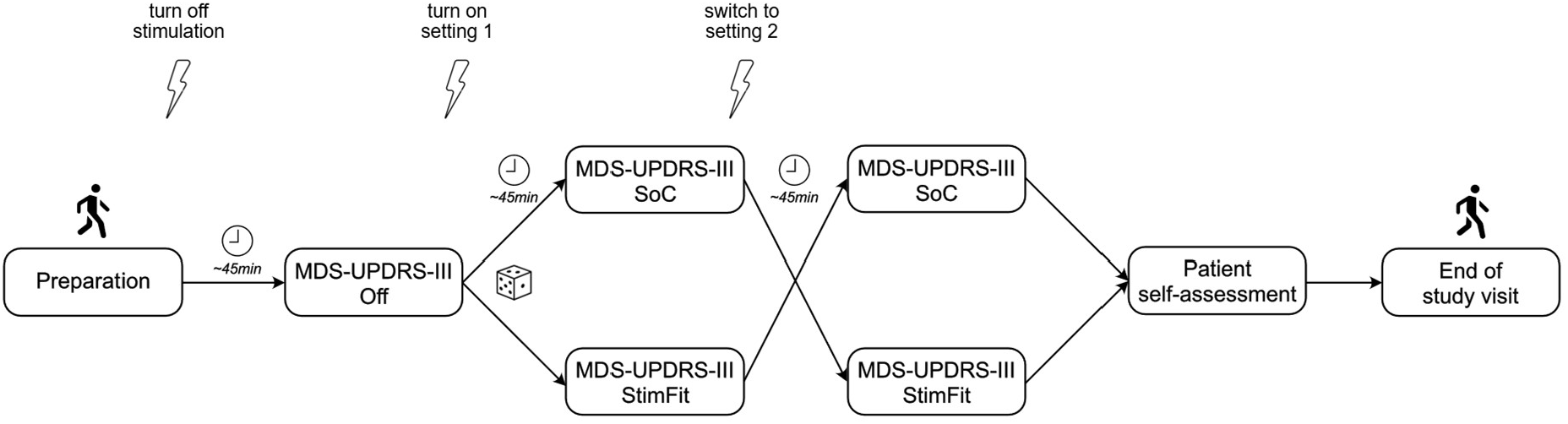
Study protocol. Patients underwent study examinations after overnight withdrawal of dopaminergic medication. In the preparation phase contact impedances were measured and *StimFit* settings were programmed and activated for a short duration to detect potential acute stimulation-induced side-effects. Amplitudes were reduced in those cases until side-effects disappeared. MDS-UPDRS-III scores were assessed in OFF-stimulation conditions after a wash-out period of 45 minutes. Afterwards ON-stimulation assessments were performed under both, *StimFit* and SoC stimulation, again after a 45-minutes wash-in. Rater and patients were blinded to the sequence of both conditions which were randomized in a 1:1 ratio. After motor assessments were completed, patients were asked to self-assess both stimulation conditions on a VAS from “0 = very unsatisfactory” to “100 = very satisfactory” and to guess the correct order of both stimulation conditions. VAS= visual analogue scale; SoC= standard of care; MDS-UPDRS-III = part III of the Movement Disorder Society-Sponsored Revision of the Unified Parkinson’s Disease Rating Scale

### Masking and randomization

Rater as well as patients were blinded to the sequence of SoC and *Stimfit* stimulation conditions. The sequence was randomized in a 1:1 ratio without applying block stratification. An unmasked research assistant (APK) generated the allocation schedule using a computerized random number generator. The same researcher activated the stimulation settings during the study visit but was otherwise not involved in patient care, clinical ratings, or data analysis.

### Outcome parameters

Demographic and treatment data at the time of study participation included age, sex, disease duration, time since DBS surgery and time since last adjustment of SoC settings. SoC stimulation data was retrieved from the device and included stimulation amplitude, pulse-width, and frequency as well as the relative distribution of electric current across contacts, contact polarities and impedances. Dates and type of in-and out-patient visits related to DBS treatment were obtained retrospectively from electronic medical records at Charité and Beelitz Hospital for Parkinson’s disease. Levodopa equivalent daily doses (LEDDs) were calculated based on pre-operative medical records as well as medication schedules at the time-point of study examination.^27,28^

Primary outcome parameter was the difference between total MDS-UPDRS-III score under SoC and *StimFit* stimulation conditions. Additionally, mean improvement of MDS-UPDRS motor score relative to OFF stimulation was calculated for both stimulation conditions to verify effective DBS. Secondary outcome parameters were differences between MDS-UPDRS-III sub-scores for akinetic-rigid, tremor and axial symptoms under both conditions. A definition of MDS-UPDRS-III items included in each of the scores is provided in the **suppl. material**. Further secondary outcome parameter was the patients’ self-rating of the subjective perception of both ON-stimulation settings. Patients were asked to rate each setting according to the question: “How satisfied were you with the overall effects of the stimulation?” on a visual analogue scale (VAS) from 0 = “very unsatisfied” to 100 = “very satisfied”. Additionally, patients were asked to guess the randomization sequence of both conditions which was documented on a binary scale as “correct” or “incorrect”. Permanent side-effects that might have occurred during initial programming of *StimFit* settings were documented along with the reduction of the stimulation amplitude necessary to achieve side-effect relieve. Post-hoc, energy efficiency of both settings was estimated according to the formulas for multiple independent current control published by Zhang et al (details described in the **suppl. material**).^23,29,30^ Patients with bipolar SoC stimulation settings were excluded from this analysis.

### Sample size calculation and statistical analysis

The study was powered to assess non-inferiority of *StimFit* compared to SoC settings using absolute differences between corresponding MDS-UPDRS-III scores as a primary endpoint. Schrag et al suggested that a difference in improvements of at least five points should be considered clinically significant.^31^ Using a one-sided t-test with an alpha of 5 % and power of 80 % a sample size of n = 35 was needed to show non-inferiority with a margin of five points. Calculations were based on an estimated mean difference between treatments of 0 ± 16.4 points, which was derived from a comparable cohort from our center.^9^ The sample-size calculation was supported by the local statistics department using the *TrialSize*-package in R.

In addition to non-inferiority, the primary outcome was tested for superiority using a two-sided t-test. Secondary endpoints included absolute differences in MDS-UPDRS-III sub-scores, patients’ VAS self-ratings and battery drain under *StimFit* and SoC stimulation and were analyzed using Wilcoxon signed rank tests. Patients were excluded from specific MDS-UPDRS-III sub-score analyses if they did not experience the corresponding symptom at baseline (sub-score in OFF-stimulation state equal to zero). A two-sided binomial test was applied to evaluate whether patients could guess the correct order of stimulation settings above chance level. Significance levels were set to an α of 0.05, and no multiplicity adjustment was applied. Statistical analyses were conducted using Matlab 2020a.

## RESULTS

Medical records of 86 PD patients who underwent bilateral STN-DBS surgery at Charité between Apr 1, 2018 and Apr 30, 2021 were screened for eligibility criteria. Out of the remaining 67 potential study participants, 52 patients were contacted for recruitment until the intended sample size of n = 35 (32 Boston Vercise™ Cartesia, 3 Medtronic SenSight™) was achieved (**Supp. Fig. 2**). Study visits were conducted between Oct 23, 2020 and Oct 28, 2021. All patients who participated in the study were included in the analysis. The first four patients were reinvited for study participation after a final patch to the *StimFit* software was applied and data from their first study visits were excluded from the analysis. Patient demographics and treatment data as well as electrode localizations are summarized in the supplementary material (**Suppl.Table 1, Suppl. Fig. 3**). OFF-stimulation MDS-UPDRS-III scores improved from 47.3 ± 17.1 to 24.7 ± 12.4 (48 %) and 26.3 ± 12.4 (43 %) under SoC and *StimFit* stimulation, respectively (**Fig. 3A**). Mean difference between motor scores was -1.6 ± 7.1 (95% CI: [-4.0, 0.9], p = 0.20, n = 35) establishing non-inferiority at a margin of five points (p = 0.0038, **Fig. 3B**). 17 patients (49 %) had better motor scores under *StimFit* stimulation compared to SoC.

**Figure 3:**
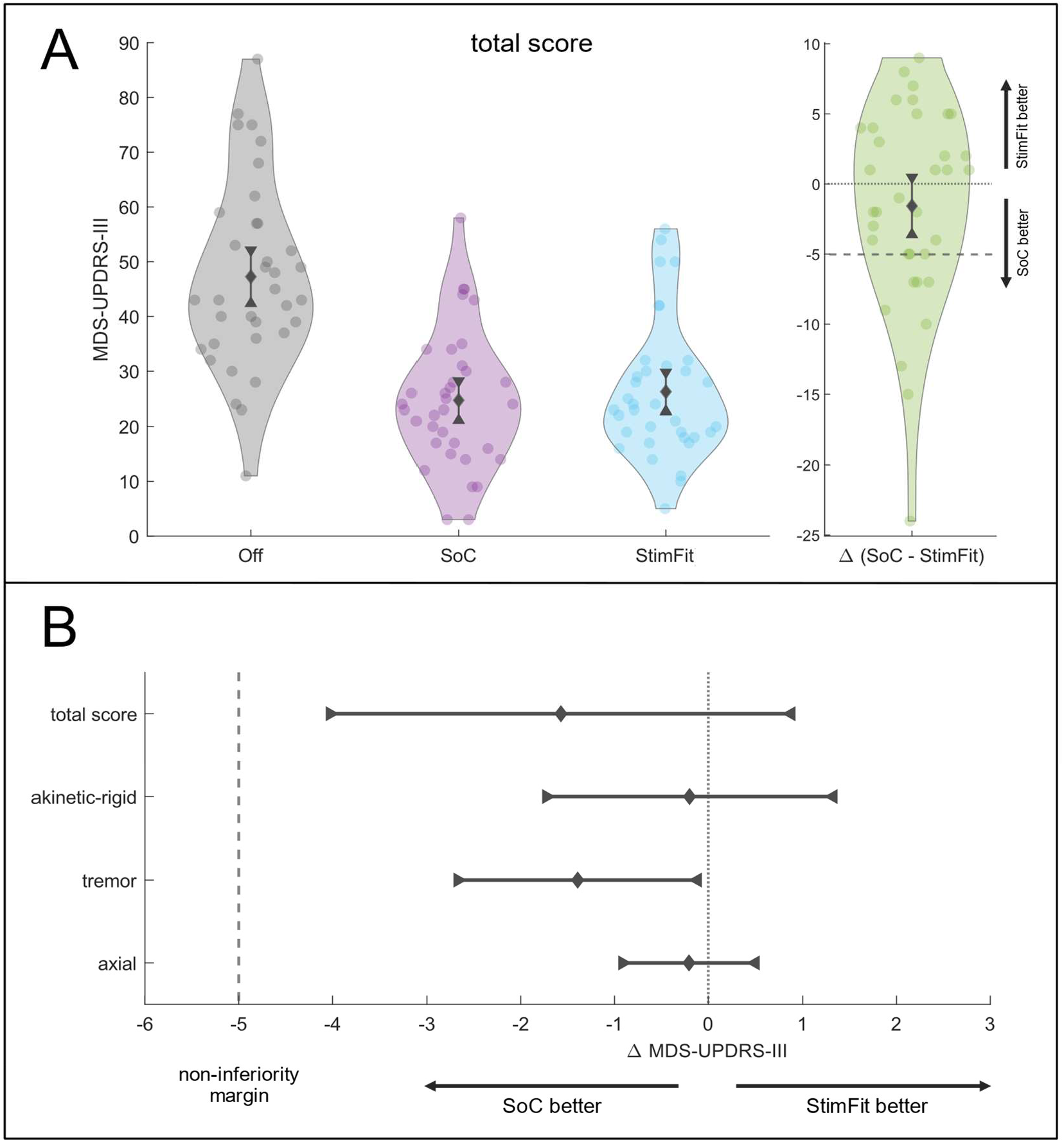
Primary endpoint and summary statistics. **A:** Violin plots showing total MDS-UPDRS-III scores under OFF (grey), SoC (purple) and *StimFit* (blue) stimulation conditions on the left as well as differences of motor scores between StimFit and SoC stimulation on the right (green). Mean and 95% CIs are displayed at each plot. **B:** Summary statistics of the primary endpoint and symptom-specific MDS-UPDRS-III sub-scores. Mean absolute differences between both ON-stimulation conditions are shown together with their 95% CIs. The CI of the total score did not include the margin of five points, establishing non-inferiority. SoC= standard of care; CI = confidence interval; MDS-UPDRS-III = part III of the Unified Parkinson’s Disease Rating Scale

Analysis of MDS-UPDRS-III sub-scores revealed an improvement for akinesia-rigidity of 46 % and 43 %, tremor 70 % and 62 %, and axial score 36 % and 35 % under SoC and *StimFit* stimulation, respectively **(Fig. 4)**. This resulted in following mean differences in sub-scores between both stimulation conditions (**Fig. 3B**): Akinesia-rigidity score: -0.2 ± 4.4 (95% CI: [-1.7, 1.3], p =0.98, n = 35), tremor score: -1.4 ± 3.3, (95% CI: [-2.7, -0.1], p = 0.046, n= 28) and axial score: -0.2 ± 2.0 (95% CI: [-0.9, 0.5], p = 0.67, n = 34).

**Figure 4:**
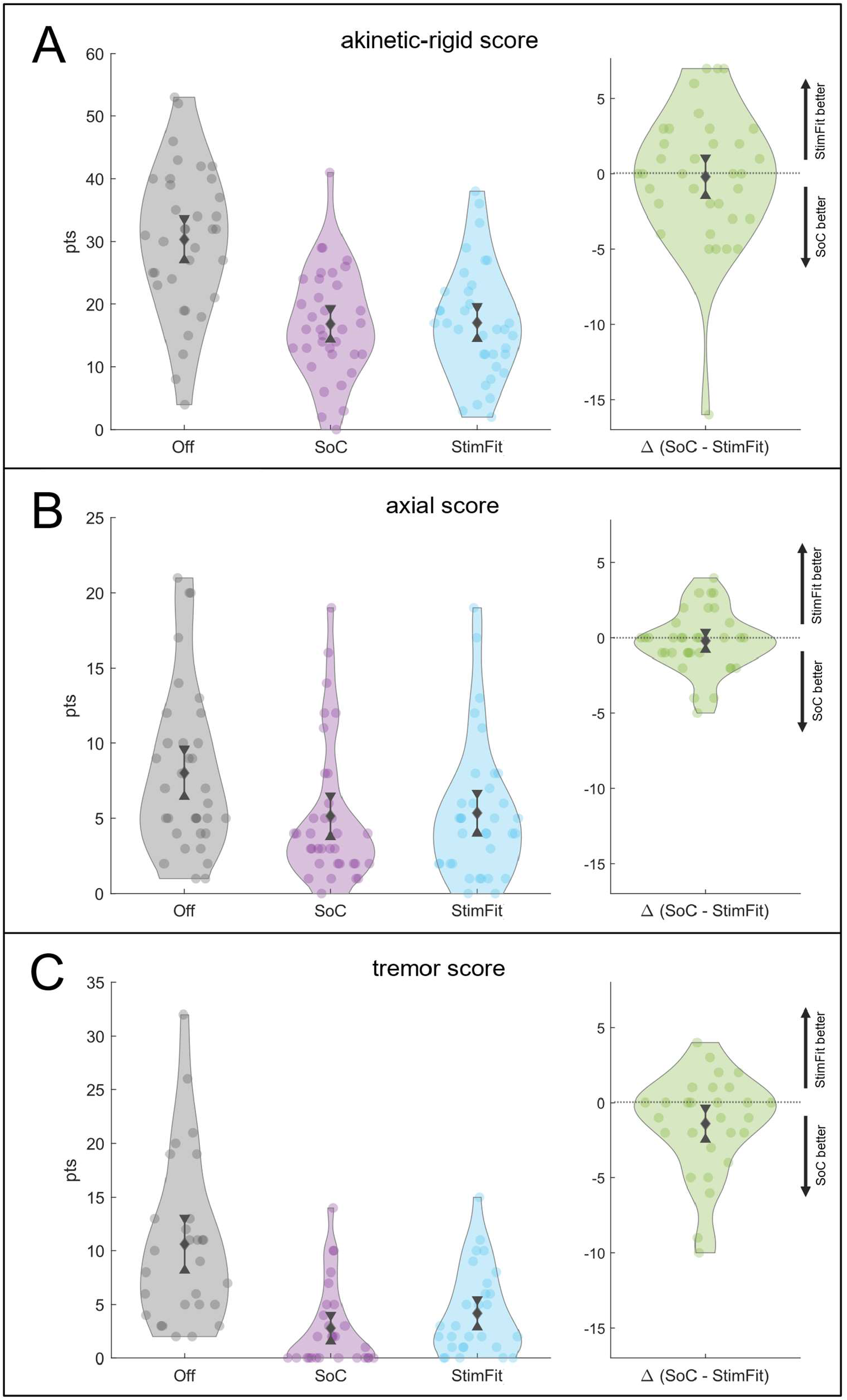
Secondary endpoint I: Symptom-specific motor effects. Violin plots showing symptom specific motor scores for akinetic-rigid **(A)** and axial **(B)** symptoms, as well as tremor **(C)**. Each panel depicts scores under OFF (grey), SoC (purple) and *StimFit* (light blue) stimulation conditions on the left and differences of motor scores between StimFit and SoC stimulation on the right (green). Mean and 95% CIs are displayed at each plot. SoC= standard of care; CI = confidence interval

Patients’ self-assessments of the overall stimulation effects from 0 = “very unsatisfactory” to 100 = “very satisfactory” showed significant differences in favor of SoC, with SoC settings on average scoring 74 ± 19 points as compared to 55 ± 24 under *StimFit* stimulation (mean difference: 19 ± 28, 95% CI: [9, 29], p < 0.001, n = 34, **Fig. 5**). *StimFit* settings were rated superior to SoC by eight (24 %) and equal ratings were given by three patients (9 %). 19 patients (56 %) correctly guessed the order of stimulation conditions, which was not significantly above chance level (p = 0.50). One patient was accidentally unblinded before the first ON-stimulation assessment by looking at the patient programmer and was therefore excluded from the self-assessment analyses. The patient was not excluded from the remaining analyses since the sequence was not revealed to the rater before the study visit was completed.

**Figure 5:**
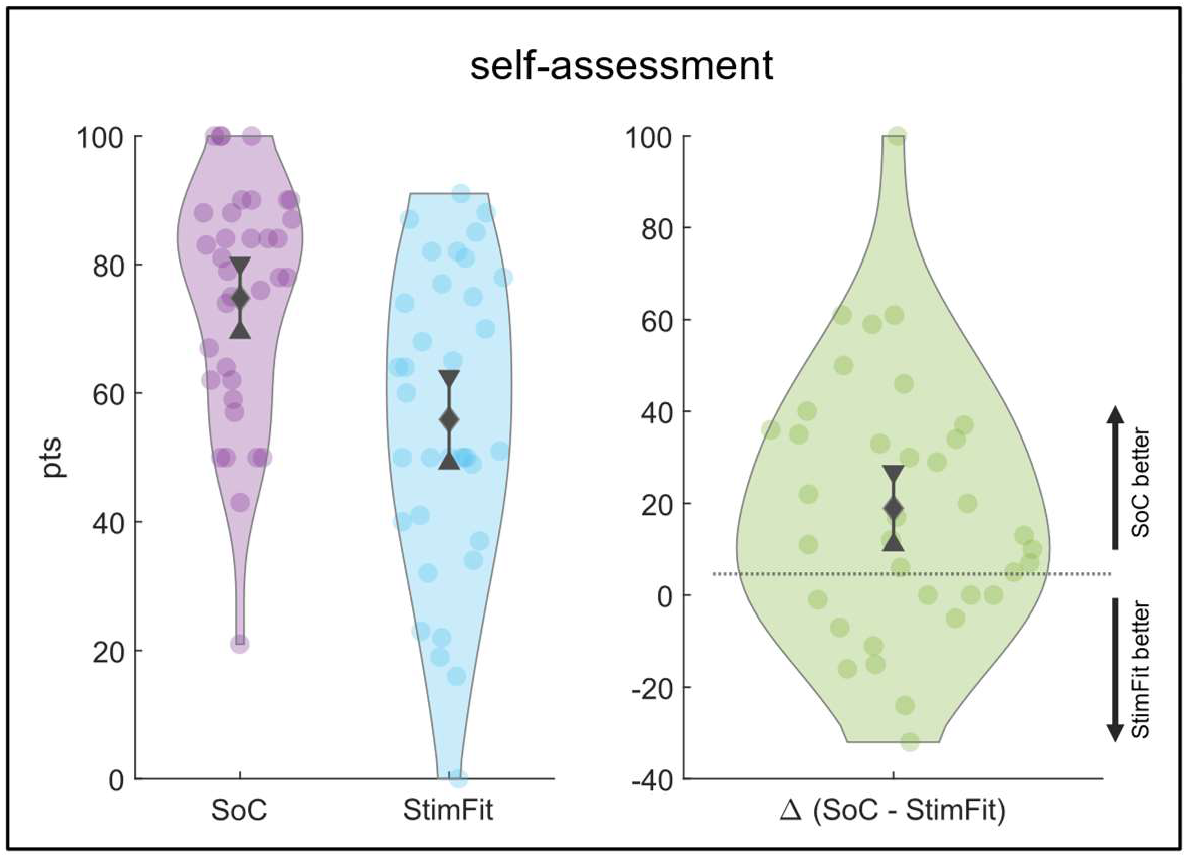
Secondary outcome II: Self-assessments. Violin plots showing results of patient ratings of both SoC (purple) and StimFit (light blue) stimulation conditions on the left as well as differences between ratings on the right (green). Mean and 95% CIs are displayed at each plot. SoC= standard of care; CI = confidence interval

In six patients (17 %) initial programming of *StimFit* settings (during the preparation phase) resulted in acute side-effects (two muscle contractions, two dysarthria, two vertigo). Stimulation amplitudes were reduced until side-effects disappeared (mean reduction 0.41 ± 0.16 mA ranging from 0.3 to 0.7 mA). Delayed onset dyskinesias appeared in three patients under *Stim-Fit* stimulation. In two cases these were rated as severe and potentially interfered with motor ratings. Dyskinesias were also observed in two cases under SoC stimulation but did not affect motor assessments.

LEDDs were reduced by 57 % under SoC treatment compared to pre-surgical medication and SoC settings had remained unchanged for 6.7 ± 5.8 months prior to study participation. Mean stimulation amplitudes were 2.6 ± 1.1 mA and an average of 3.2 ± 1.4 contacts were active. Bipolar settings were applied on four electrodes. One patient was treated with interleaving stimulation bilaterally. Mean pulse-widths and frequencies were 60 ± 3 μs and 134 ± 25 Hz. Mean stimulation amplitude suggested by *StimFit* was 2.4 ± 0.3 mA distributed across 5.4 ± 2.1 active contacts. Battery drain was estimated to be 57 ± 29 μA in SoC compared to 50 ± 21 μA in *StimFit* settings (p = 0.5, n = 32). All impedance values were within normal ranges. The parameters of both stimulation conditions are provided in the supplementary material for each subject and in a summarized form (**Supplementary Tables 2 & 3**).

Since SoC treatment was conducted before study recruitment the exact times spent for parameter optimization in each patient were not available. However, during 20 ± 11 months since DBS surgery, patients received DBS-related treatment at specialized in-and out-patient facilities on 28 ± 16 days, including thorough monopolar reviews and OFF-medication assessments. The *StimFit* algorithm converged to a final solution within ∼50 minutes in all patients and did not require manual steps.

## DISCUSSION

In this prospective double-blind cross-over randomized non-inferiority trial bilateral STN-DBS was applied in 35 PD patients using stimulation parameters suggested by an automated algorithm (*StimFit*) based on electrode locations. *StimFit* stimulation reduced motor symptoms with a statistically non-significant mean difference of 1.6 points compared to SoC on the MDS-UPDRS-III scale, establishing non-inferiority within a predefined margin of five points. Importantly, both stimulation conditions resulted in significant motor improvements of 48 % and 43 % compared to OFF-stimulation baseline confirming effective STN-DBS and sufficient wash-out/wash-in time between conditions in this patient group.^1-3^ In line with this, previous long-term SoC treatment had led to reduction of dopaminergic medication (LEDD) by 57% compared to preoperative treatment in our cohort.

A strength of this study was the prospective double-blind cross-over design and the fact that it used for the first time a data-driven algorithm capable of suggesting optimal stimulation parameters in PD patients treated with STN-DBS based on electrode location in a fully automated fashion. A limitation was the short observational period, which precluded assessment of long-term motor effects and quality of life. Moreover, only cathodal contact configurations are suggested since the algorithm has not been tested for bipolar or interleaving settings. In addition, the algorithm was trained and validated on data with a fixed pulse-width of 60 μs and stimulation frequency of 130 Hz. It does not allow to account for effects of varying frequencies, pulse-shapes or -durations. We further noticed that the variation of stimulation amplitudes suggested by *StimFit* was relatively small compared to SoC. Initial amplitudes ranged between 2.3 and 2.5 mA across the cohort at a side-effect threshold of 20 %. This might indicate that variance in electrode location and active contact configuration only had limited impact on side-effect predictions compared to stimulation amplitude and suggests that our modeling approach for side-effects should be optimized further.

Ever more complex DBS electrode designs call for guided programming by integrating biomarkers to identify beneficial combinations of stimulation parameters. Computational models of different complexities are available linking the relationship between electrode location, stimulation parameters and clinical outcome.^32-38^ Due to its simplicity and low computational demands the binarized approach of modeling the volume of activated tissue (VTA) in respect to patients’ brain anatomy has become a popular method and was recently implemented in commercially available software.^19^ However, “bottom-up” biophysical models of neuronal activation are based on many pre-assumptions, most of which are unknown in the individual patient, e.g., fiber diameters and their orientation relative to the electric field, which has shown to impact fiber activation *in silico* and *in vivo*.^39,40^ Further, direct axonal activation is the only mechanism of action considered by the model neglecting potential stimulation effects on dendrites, glial cells, intracellular cascades, transmitter depletion and network dynamics.^41^ Consequently, the VTA should be considered a rather vague metric to estimate the anatomical regions affected by stimulation. To complicate things even more, exact target regions, as well as regions of avoidance are still a matter of debate and were not defined within currently available software for image-guided DBS. This requires physicians to adjust DBS parameters in respect to visible anatomical structures without a clear optimization objective leaving room for different solutions based on the anatomical intuition and training of the programmer. Finally, this *in silico* optimization needs to be performed manually demanding time and resources. With *StimFit* we aimed at overcoming some of these limitations. First, the underlying prediction model is based on the properties of the electric field in the vicinity of the electrode and does not require assumptions about mechanisms of action of DBS. Predictions made by this biophysically naïve approach could proof to be more robust across different behavioral effects since they are independent of the underlying neurophysiological mechanisms.

Second, the model was trained and cross-validated on a large sample of monopolar review data allowing to predict and quantify stimulation effects. Predictions were tested on an independent test dataset before, third, the model was embedded in a mathematical optimization framework to identify optimal stimulation settings in a time-efficient manner and without need of any manual interactions.

While many studies in the field of DBS neuroimaging have applied out-of-sample testing to retrospectively validate their results ^10,11,13-15^, prospective applications have remained sparse. Frankemolle et al. used image-based models to identify stimulation settings which would minimize spread of electric current to nonmotor regions of the STN.^42^ Applied in ten patients, these settings led to a better performance in a working memory task compared to standard of care stimulation and they concluded that cognitive decline associated with STN-DBS could be avoided by using model-based stimulation parameters. Other prospective studies concluded that software-assisted programming was significantly faster compared to standard clinical procedures.^20-22^ In three studies, model-based parameters achieved therapeutic benefit comparable to clinical programming after parameters were re-adjusted based on behavioral feedback. No adjustments were made in a study by Waldthaler et al. and non-inferiority was reported at a margin of 20 % improvement relative to OFF-stimulation baseline. However, keeping in mind that larger controlled DBS trials have reported mean motor improvements ranging from 24 % to 49 %, this margin should be considered too liberal.^43^

Interestingly, in our cohort akinetic-rigid and axial sub-scores showed similar improvements under both stimulation conditions, while tremor responded significantly less to *StimFit* stimulation. Our understanding of anatomical target structures involved in therapeutic neuromodulation is currently undergoing a paradigm shift from a disease-to a symptom-centric view.^44^ More specifically, in PD, multiple recent publications point toward an anatomical segregation of DBS “sweetspots” for suppression of tremor on the one hand and akinesia and rigidity on the other hand.^13^ Moreover, the dentato-rubro-thalamic tract might be a common target structure for the suppression of tremor across different pathologies.^45^ This has potential implications for personalized DBS programming procedures since optimal DBS parameters would depend on individual patients’ symptom profiles. Within the *StimFit* software this concept is now implemented by allowing to modify the optimization objective (motor symptom control) to maximize the predicted therapeutic effects of tremor or akinetic-rigid symptoms on a continuous spectrum. In this present study, however, we did not yet include tremor subitems from *StimFit* predictions, forcing the model to find settings which would maximize improvement of akinetic-rigid symptoms, exclusively. Seeing this being reflected in our sub-score analysis provides prospective evidence to support the assumption of symptom-specific stimulation sites and underlines the importance of taking baseline symptomatology into account in automated DBS programming procedures. However, further prospective studies are necessary to proof that optimization procedures adapted to patients’ symptom profile will yield higher clinical benefit.

Despite non-inferiority of *StimFit* stimulation being established for objective motor assessments, subjective patient ratings showed significant differences favoring SoC. To investigate potential reasons for this discrepancy we explored the characteristics of patients which clearly preferred SoC ratings over *StimFit*. We found that the presence of hyperkinetic symptoms under *StimFit* stimulation was associated with lower VAS ratings compared to SoC. More specifically, patients who experienced suboptimal tremor control or strong dyskinesias under *StimFit* stimulation strongly favored SoC settings. This led us to conclude that hyperkinetic symptoms might have excessively contributed to VAS ratings due to the explicitly observable patient feedback. An optimization procedure tailored to patients’ symptomatic profile might therefore also improve their subjective perception of treatment benefit.

Stimulation settings differed between *StimFit* and SoC with respect to numbers of contacts activated with more multi-contact configurations suggested by the algorithm. Clinical optimization strategies usually start by conducting a monopolar review to assess therapeutic windows of stimulation amplitudes for individual contacts or contact levels. Based on these information, more complex multi-contact configurations which might provide superior benefit will then be probed. This process is carried out analogously by *StimFit* which starts the optimization procedure by predicting motor and side-effects at different amplitudes to identify favorable monopolar solutions which are then used as starting points for a gradient descent algorithm to explore potentially superior multi-cathode solutions. The number of iterations in this optimization procedure depends on the solver’s stopping criteria, such as the changes in predicted benefit in previous iterations. Clinical programming strategies also need to apply certain stopping criteria, but due to the limited number of iterations those need to be much more liberal, so that further adjustments are often only made when therapeutic outcome is clearly suboptimal, or patients are not satisfied with treatment effects. In many cases this results in monopolar, or pseudo-monopolar stimulation settings (equal distribution of electric current across all contacts at one segmented level), which were chosen for SoC in 46 % of the cases in this cohort. *StimFit*, however, predicted superior clinical outcome for multi-contact configurations in 93 % of the cases. Overall, *StimFit* solutions show a more gradual distribution of electric current across contacts compared to SoC. Such settings could not be derived from clinical trial-and-error programming but might bear potential therapeutic advantages, underlining the importance of automated optimization strategies in modern DBS devices. Importantly, battery drain was similar for both stimulation settings in our study. This was in contrast to a reduction in estimated battery drain that has been reported for stimulation settings obtained from anatomy-guided programming.^23^ Future algorithms could incorporate estimated energy efficiency as an additional variable to obtain settings with an optimized battery life cycle.

In conclusion, results of this prospective randomized double-blind cross-over trial showed that application of STN-DBS parameters suggested by a data-driven optimization algorithm in a cohort of 35 PD patients led to a significant reduction of motor impairment compared to OFF stimulation, similar to the effects obtained during SoC stimulation. This suggests that data-driven strategies which allow for quantitative predictions of stimulation effects, embedded in mathematical optimization procedures, could govern future programming strategies. This could not only reduce programming time and resources but might pave the way for novel electrode designs to further optimize treatment benefit.

## Data Availability

Behavioral data as well as data on stimulation settings are provided in the supplementary material. Other data supporting the findings of this study are available upon reasonable request from the corresponding author but will not be made publicly available due to their containing information that could compromise the privacy of study participants.

## CONTRIBUTORS

JR, TAD, AH and AAK conceptualized the study. Funding was acquired by JR and AAK. JR, JA, JB, APK, GHS and PK contributed to data acquisition. JR conducted the statistical analysis and data visualization. JA and JB verified the data and reviewed the analysis. TAD and AAK contributed to data interpretation. AAK supervised and administered the study. JR wrote the first draft and all other authors reviewed and commented on the report. All authors approved the final published version and agree to be accountable for all aspects of the work in ensuring that questions related to the accuracy or integrity of any part of the work are appropriately investigated and resolved.

## ACKNOWLEDGEMENTS

We thank Friederike Borngräber and Gregor Wenzel for their support during patient eligibility screening. We further thank the NeuroCure Clinical Research Center which provided funding as well as structural and administrative support (Germany’s Excellence Strategy – EXC-2049 – 390688087).

## DATA AVAILABILITY

Behavioral data as well as data on stimulation settings are provided in the **supplementary material**. Other data supporting the findings of this study are available upon reasonable request from the corresponding author but will not be made publicly available due to their containing information that could compromise the privacy of study participants.

## REFERENCES

1. Deuschl G, Schade-Brittinger C, Krack P, et al. A randomized trial of deep-brain stimulation for Parkinson’s disease. N Engl J Med 2006;355:896–908.

2. Deep-Brain Stimulation for Parkinson’s Disease Study G, Obeso JA, Olanow CW, et al. Deep-brain stimulation of the subthalamic nucleus or the pars interna of the globus pallidus in Parkinson’s disease. N Engl J Med 2001;345:956–63.

3. Limousin P, Krack P, Pollak P, et al. Electrical stimulation of the subthalamic nucleus in advanced Parkinson’s disease. N Engl J Med 1998;339:1105–11.

4. Okun MS, Tagliati M, Pourfar M, et al. Management of referred deep brain stimulation failures: a retrospective analysis from 2 movement disorders centers. Arch Neurol 2005;62:1250–5.

5. Volkmann J, Moro E, Pahwa R. Basic algorithms for the programming of deep brain stimulation in Parkinson’s disease. Mov Disord 2006;21 Suppl 14:S284–9.

6. Moro E, Esselink RJ, Xie J, Hommel M, Benabid AL, Pollak P. The impact on Parkinson’s disease of electrical parameter settings in STN stimulation. Neurology 2002;59:706–13.

7. Dembek TA, Reker P, Visser-Vandewalle V, et al. Directional DBS increases side-effect thresholds-A prospective, double-blind trial. Mov Disord 2017;32:1380–8.

8. Connolly MJ, Cole ER, Isbaine F, et al. Multi-objective data-driven optimization for improving deep brain stimulation in Parkinson’s disease. J Neural Eng 2021;18.

9. Wenzel GR, Roediger J, Brucke C, et al. CLOVER-DBS: Algorithm-Guided Deep Brain Stimulation-Programming Based on External Sensor Feedback Evaluated in a Prospective, Randomized, Crossover, Double-Blind, Two-Center Study. J Parkinsons Dis 2021.

10. Al-Fatly B, Ewert S, Kubler D, Kroneberg D, Horn A, Kuhn AA. Connectivity profile of thalamic deep brain stimulation to effectively treat essential tremor. Brain 2019;142:3086–98.

11. Baldermann JC, Melzer C, Zapf A, et al. Connectivity Profile Predictive of Effective Deep Brain Stimulation in Obsessive-Compulsive Disorder. Biol Psychiatry 2019.

12. Dembek TA, Barbe MT, Astrom M, et al. Probabilistic mapping of deep brain stimulation effects in essential tremor. Neuroimage Clin 2017;13:164–73.

13. Dembek TA, Roediger J, Horn A, et al. Probabilistic sweet spots predict motor outcome for deep brain stimulation in Parkinson disease. Ann Neurol 2019;86:527–38.

14. Horn A, Reich M, Vorwerk J, et al. Connectivity Predicts deep brain stimulation outcome in Parkinson disease. Ann Neurol 2017;82:67–78.

15. Reich MM, Horn A, Lange F, et al. Probabilistic mapping of the antidystonic effect of pallidal neurostimulation: a multicentre imaging study. Brain 2019.

16. Horn A, Kuhn AA. Lead-DBS: a toolbox for deep brain stimulation electrode localizations and visualizations. Neuroimage 2015;107:127–35.

17. Horn A, Li N, Dembek TA, et al. Lead-DBS v2: Towards a comprehensive pipeline for deep brain stimulation imaging. Neuroimage 2019;184:293–316.

18. Lauro PM, Vanegas-Arroyave N, Huang L, et al. DBSproc: An open source process for DBS electrode localization and tractographic analysis. Hum Brain Mapp 2016;37:422–33.

19. Miocinovic S, Noecker AM, Maks CB, Butson CR, McIntyre CC. Cicerone: stereotactic neurophysiological recording and deep brain stimulation electrode placement software system. Acta Neurochir Suppl 2007;97:561–7.

20. Pavese N, Tai YF, Yousif N, Nandi D, Bain PG. Traditional Trial and Error versus Neuroanatomic 3-Dimensional Image Software-Assisted Deep Brain Stimulation Programming in Patients with Parkinson Disease. World Neurosurg 2020;134:e98–e102.

21. 1. Pourfar MH, Mogilner AY, Farris S, et al. Model-Based Deep Brain Stimulation Programming for Parkinson’s Disease: The GUIDE Pilot Study. Stereotact Funct Neurosurg 2015;93:231–9.

22. Waldthaler J, Bopp M, Kuhn N, et al. Imaging-based programming of subthalamic nucleus deep brain stimulation in Parkinson’s disease. Brain Stimul 2021;14:1109–17.

23. Lange F, Steigerwald F, Malzacher T, et al. Reduced Programming Time and Strong Symptom Control Even in Chronic Course Through Imaging-Based DBS Programming. Front Neurol 2021;12:785529.

24. Noecker AM, Frankemolle-Gilbert AM, Howell B, et al. StimVision v2: Examples and Applications in Subthalamic Deep Brain Stimulation for Parkinson’s Disease. Neuromodulation 2021;24:248–58.

25. Roediger J, Dembek TA, Wenzel G, Butenko K, Kuhn AA, Horn A. StimFit-A Data-Driven Algorithm for Automated Deep Brain Stimulation Programming. Mov Disord 2021.

26. Picillo M, Lozano AM, Kou N, Puppi Munhoz R, Fasano A. Programming Deep Brain Stimulation for Parkinson’s Disease: The Toronto Western Hospital Algorithms. Brain Stimul 2016;9:425–37.

27. Schade S, Mollenhauer B, Trenkwalder C. Levodopa Equivalent Dose Conversion Factors: An Updated Proposal Including Opicapone and Safinamide. Mov Disord Clin Pract 2020;7:343–5.

28. Tomlinson CL, Stowe R, Patel S, Rick C, Gray R, Clarke CE. Systematic review of levodopa dose equivalency reporting in Parkinson’s disease. Mov Disord 2010;25:2649–53.

29. Juarez-Paz LM. In silico Accuracy and Energy Efficiency of Two Steering Paradigms in Directional Deep Brain Stimulation. Front Neurol 2020;11:593798.

30. Zhang S, Silburn P, Pouratian N, et al. Comparing Current Steering Technologies for Directional Deep Brain Stimulation Using a Computational Model That Incorporates Heterogeneous Tissue Properties. Neuromodulation 2020;23:469–77.

31. Schrag A, Sampaio C, Counsell N, Poewe W. Minimal clinically important change on the unified Parkinson’s disease rating scale. Mov Disord 2006;21:1200–7.

32. Astrom M, Diczfalusy E, Martens H, Wardell K. Relationship between neural activation and electric field distribution during deep brain stimulation. IEEE Trans Biomed Eng 2015;62:664–72.

33. Butson CR, Cooper SE, Henderson JM, McIntyre CC. Patient-specific analysis of the volume of tissue activated during deep brain stimulation. Neuroimage 2007;34:661–70.

34. Duffley G, Anderson DN, Vorwerk J, Dorval AD, Butson CR. Evaluation of methodologies for computing the deep brain stimulation volume of tissue activated. J Neural Eng 2019;16:066024.

35. Frankemolle-Gilbert AM, Howell B, Bower KL, Veltink PH, Heida T, McIntyre CC. Comparison of methodologies for modeling directional deep brain stimulation electrodes. PLoS One 2021;16:e0260162.

36. Gunalan K, Chaturvedi A, Howell B, et al. Creating and parameterizing patient-specific deep brain stimulation pathway-activation models using the hyperdirect pathway as an example. PLoS One 2017;12:e0176132.

37. Howell B, Gunalan K, McIntyre CC. A Driving-Force Predictor for Estimating Pathway Activation in Patient-Specific Models of Deep Brain Stimulation. Neuromodulation 2019;22:403–15.

38. Maks CB, Butson CR, Walter BL, Vitek JL, McIntyre CC. Deep brain stimulation activation volumes and their association with neurophysiological mapping and therapeutic outcomes. J Neurol Neurosurg Psychiatry 2009;80:659–66.

39. Anderson DN, Duffley G, Vorwerk J, Dorval AD, Butson CR. Anodic stimulation misunderstood: preferential activation of fiber orientations with anodic waveforms in deep brain stimulation. J Neural Eng 2019;16:016026.

40. Lehto LJ, Slopsema JP, Johnson MD, et al. Orientation selective deep brain stimulation. J Neural Eng 2017;14:016016.

41. 1. Jakobs M, Fomenko A, Lozano AM, Kiening KL. Cellular, molecular, and clinical mechanisms of action of deep brain stimulation-a systematic review on established indications and outlook on future developments. EMBO Mol Med 2019;11.

42. Frankemolle AM, Wu J, Noecker AM, et al. Reversing cognitive-motor impairments in Parkinson’s disease patients using a computational modelling approach to deep brain stimulation programming. Brain 2010;133:746–61.

43. Krack P, Volkmann J, Tinkhauser G, Deuschl G. Deep Brain Stimulation in Movement Disorders: From Experimental Surgery to Evidence-Based Therapy. Mov Disord 2019;34:1795–810.

44. Hollunder B, Rajamani N, Siddiqi SH, et al. Toward personalized medicine in connectomic deep brain stimulation. Prog Neurobiol 2021;210:102211.

45. Listik C, Santiago N, Reis PR, et al. Targeting the hot spot in a patient with essential tremor and Parkinson’s disease: Tractography matters. Clin Neurol Neurosurg 2018;174:230–2.

