## Supplementary Material for "Automated Deep Brain Stimulation programming based on electrode location – a randomized, cross-over trial using a data-driven algorithm"

### Energy efficiency

Current draw from the battery was calculated according to previously published equations for multiple independent current control (MICC) [1,2]:

$$I_{MICC} = I_{overhead} + \sum_{i=1}^{N} I_{Ei} * PW * f * \frac{V_{max}}{V_{bat}}$$

Where:

$I_{MICC}$: Current draw from battery
$I_{overhead}$: Frequency-dependent IPG overhead current, which was set to 4.9 µA [2]

N: Number of activated contacts

$I_{Ei}$: Pulse amplitude for contact i

PW: Pulse width

f: Pulse frequency

$V_{max}$: Maximum voltage for the activated electrodes

$V_{bat}$: Battery voltage, which was set to 2.8 V [2]

And

$$V_{max} = \max\{(I_{Ei} * Z_{Ei}) : i = 1..N\}$$

Where:

$V_{max}$: Maximum voltage for the activated contacts

$I_{Ei}$: Pulse amplitude for contact i

$Z_{Ei}$: Impedance of contact i

N: Number of activated contacts

### Symptom-specific sub-scores

Following items of the MDS-UPDRS-III were included in the symptom-specific analysis:

- Akinetic-rigid score: Items 3.2 to 3.8
- Axial score: Items 3.9 to 3.14
- Tremor score: 3.15 to 3.18

### References


1. Zhang S, Silburn P, Pouratian N, et al. Comparing Current Steering Technologies for Directional Deep Brain Stimulation Using a Computational Model That Incorporates Heterogeneous Tissue Properties. Neuromodulation 2020;23:469-77.
2. Juarez-Paz LM. In silico Accuracy and Energy Efficiency of Two Steering Paradigms in Directional Deep Brain Stimulation. Front Neurol 2020;11:593798.
