## Supplementary Figures for "Automated Deep Brain Stimulation programming based on electrode location – a randomized, cross-over trial using a data-driven algorithm"

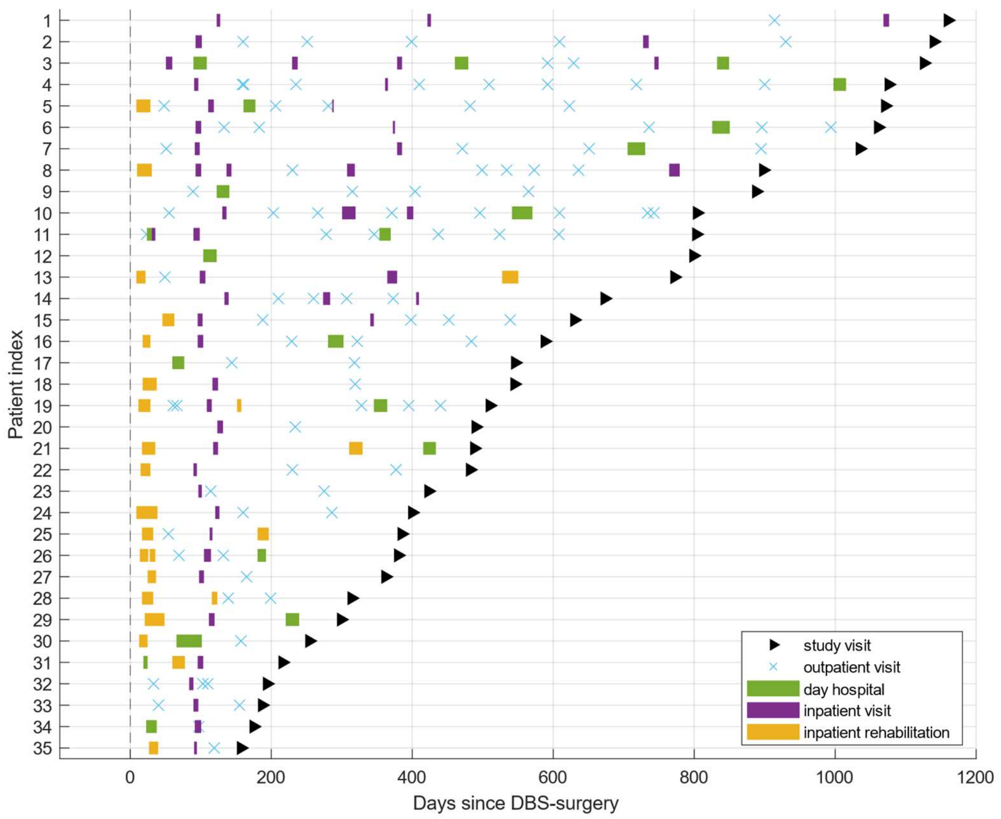


***Supplementary Figure 1: DBS-related in- and outpatient treatment since surgery***

*Follow-up in- and outpatient treatment related to clinical optimization of DBS parameters between DBS-surgery and study visit. Data were obtained retrospectively from electronic medical records at Charité University medicine and Beelitz Hospital for neurological early rehabilitation. To protect patient confidentiality, patient indices were not aligned with those reported in Supplementary Tables 3 and 4 but were instead sorted by duration between surgeries and study visits.*

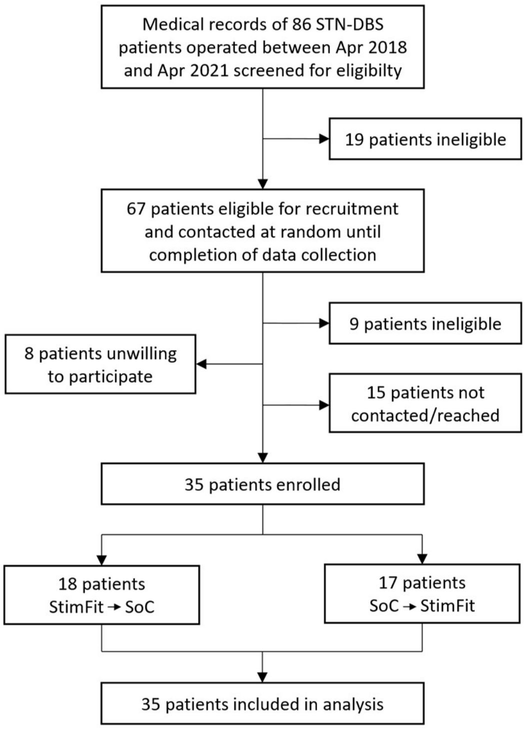


***Supplementary Figure 2: Trial profile***

*STN = subthalamic nucleus; DBS = deep brain stimulation; SoC = standard of care*

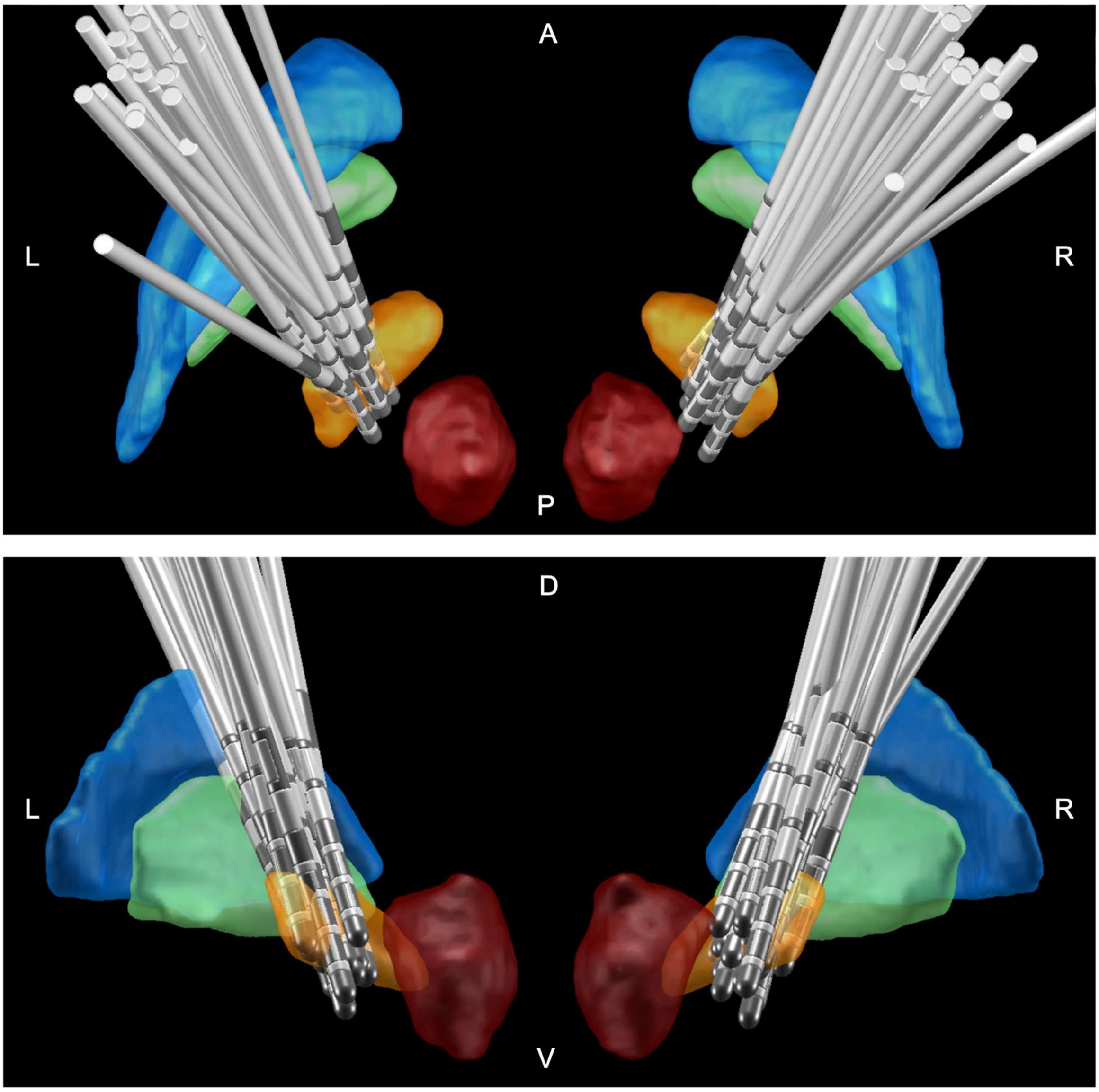


***Supplementary Figure 3:***

*Electrode localizations in MNI-space shown from dorsal (upper panel) and posterior (lower panel) in relation to the internal (green) and external (blue) parts of the globus pallidus and subthalamic (orange) and red (red) nuclei. L = left; R = right; A = anterior; P = posterior; D = dorsal; V = ventral*
